# Prodromal Lewy body disorder features in REM sleep behavior disorder with biomarker-defined synucleinopathy

**DOI:** 10.64898/2025.12.24.25342976

**Authors:** Daniel Weintraub, Michele K. York, Roseanne Dobkin, Anuprita R Nair, Ryan Kurth, David-Erick Lafontant, Chelsea Caspell-Garcia, Roy N. Alcalay, Ethan G. Brown, Lana M. Chahine, Christopher Coffey, Tatiana Foroud, Douglas Galasko, Karl Kieburtz, Kenneth Marek, Kalpana Merchant, Brit Mollenhauer, Kathleen L. Poston, Andrew Siderowf, Cristina Simonet, Tanya Simuni, Caroline M. Tanner, Thomas F. Tropea, Aleksandar Videnovic, Parkinson’s Progression Markers Initiative

## Abstract

**Objective:** Isolated rapid eye movement sleep behavior disorder (iRBD) is a prodromal state for Lewy body disorders and exhibits biological heterogeneity that may influence clinical expression and progression. We examined clinical features in individuals with iRBD and biomarker-defined synucleinopathy.

**Methods:** Parkinson’s Progression Markers Initiative (PPMI) is a longitudinal, multi-center observational study. Participants included polysomnogram (PSG)-confirmed iRBD individuals who were cerebrospinal fluid (CSF) α-synuclein seed amplification assay positive with no clinical diagnosis of Parkinson’s disease or dementia with Lewy bodies, along with robust healthy controls (HCs). Clinical and biological features of prodromal PD and DLB, including mild cognitive impairment (MCI), subthreshold parkinsonism, and a range of neuropsychiatric, autonomic and sensory symptoms, were assessed.

**Results:** Compared with HCs (*N*=136), iRBD participants (*N*=197) demonstrated worse cognitive performance, including a lower cognitive summary score (*p*<0.0003, effect size = 0.41), and higher odds of subthreshold parkinsonism (OR = 24.5), and neuropsychiatric (OR = 3.5), autonomic (OR = 7.2) and sensory symptoms (OR = 13.2). Common features included hyposmia (75%), pain (54%), urinary problems (52%), constipation (49%), lightheadedness (40%) and anxiety (36%), whereas rates of MCI (32%), subthreshold parkinsonism (27%) and psychosis (7%) were lower. iRBD participants with abnormal dopamine transporter imaging had higher anxiety scores and antidepressant use. Although only 10% met criteria for prodromal DLB due to the requirement for MCI, most exhibited multi-domain impairment.

**Interpretation:** iRBD with synucleinopathy is associated with multi-domain clinical impairment prior to clinical neurodegenerative disease diagnosis, supporting broad clinical assessment in early biomarker-defined synuclein disease.

## INTRODUCTION

Rapid eye movement sleep behavior disorder (RBD) is present in up to 75% of Parkinson’s disease (PD) and 90% of dementia with Lewy body (DLB) patients long-term.^1^ In DLB, RBD is considered a core clinical feature,^2^ and in PD, RBD symptoms are associated with faster cognitive decline^3^ and increased risk for dementia.^4^

Isolated RBD (iRBD) has emerged as the prodromal feature with the highest specificity for future risk of diagnosis of a Lewy body disorder (LBD; i.e., synucleinopathy, either PD or DLB), with an overall conversion rate from iRBD to an overt neurodegenerative syndrome of approximately 75% over 10 or more years.^5,6^ Over the past decade, research diagnostic criteria for both prodromal PD^7,8^ and DLB^9^ have been proposed, and iRBD is prominent in both sets of criteria. In addition to a focus on iRBD, cognition,^10–12^ and parkinsonism,^13^ these criteria also incorporate psychiatric,^14,15^ autonomic,^16,17^ and sensory^15,18^ symptoms, which occur more frequently in these populations compared with controls and predict progression to a clinically-defined synucleinopathy.^11,15,16,18–20^

For RBD pathophysiology, clinical and basic science suggest that α-synuclein pathology begins in the lower brainstem where REM atonia circuits are located, then propagates rostrally to brain regions such as the substantia nigra, limbic system, and cortex. Regarding presence of biologically-defined neuronal synuclein disease (NSD), in one study 85% of participants with iRBD had a positive cerebrospinal fluid [CSF] α-synuclein seed amplification assay [SAA] test,^21^ suggesting that detectable synuclein disease occurs prior to onset of significant clinical symptoms that would lead to a LBD diagnosis. Another biological marker of synuclein disease is dopaminergic dysfunction, often measured by dopamine transporter (DAT) SPECT imaging (e.g., DaTscan). In PD, mean striatal DAT SBR is inversely correlated with increasing RBD symptom score,^22^ and the mean striatal SBR value decreases more over time in patients with RBD symptoms.^23^

The Parkinson’s Progression Markers Initiative (PPMI) is a prospective cohort study that enrolls participants with prodromal PD, including iRBD. In that cohort, progression to a clinical synucleinopathy is associated with olfaction, depression, cognition, CSF ptau:total tau ratio, cortical atrophy, and dopamine transporter (DAT) SPECT imaging binding.^24,25^

There is considerable heterogeneity in the pathophysiology of iRBD, and to date, no studies have focused only on individuals with *in vivo* characterization of synucleinopathy, who would be presumed to have the highest risk and earliest likelihood of conversion to a clinically-diagnosed synuclein neurodegenerative disorder. In this context, we present the clinical and biological characteristics of a multi-center, international cohort of individuals with PSG-confirmed iRBD and biologically-defined synucleinopathy, including presence of symptoms across multiple clinical domains reported to be affected in prodromal PD and DLB. We hypothesized that iRBD patients would be more likely to have a wide range of symptoms across multiple clinical domains, and in varying combinations, compared with robust healthy controls.

## METHODS

### Cohorts

#### Isolated RBD (N=197)

Data for this analysis were from the PPMI, an international, multicenter, prospective, cohort study.^26^ The iRBD cohort were: (1) enrolled into PPMI study post-2020; (2) PSG-confirmed RBD; (3) NSD positive at baseline based on positive CSF α-synuclein SAA test (Type 1, neuronal type; see below); (4) without a local site investigator (LSI) diagnosis of PD, dementia or other neurodegenerative disease at baseline; (5) not on any PD meds; and (6) without GBA1 Gaucher-causing variants.

#### Robust healthy controls (N=136)

The robust healthy controls (HCs) were a subset of PPMI participants previously described.^27,28^ They were: (1) enrolled into the PPMI study as a HC at any time point; (2) excluded for having possible iRBD, defined by a score ≥ 6 on the RBDSQ, self-reporting an RBD diagnosis without PSG confirmation, or self-reporting dream enactment behavior; (3) NSD negative based on CSF α-synuclein SAA tests, with any longitudinal SAA test negative too; and (4) DaTscan negative.

### Variables

#### Demographic

Demographic variables were biological sex, age at baseline visit, race, ethnicity, and number of years of formal education.

#### NSD-ISS

iRBD participants were assigned to a Neuronal alpha-Synuclein Disease Integrated Staging System (NSD-ISS) stage as previously described.^29,30^ Of relevance to this paper, Stage 2 is subtle functional impairment on the basis of motor, cognitive or other non-motor symptoms; Stage 3 is slight impairment on the basis of motor or cognitive symptoms; and Stage 4 is mild impairment on the basis of motor, cognitive or other non-motor symptoms.

#### Cognitive

The LSI completed a clinical features checklist including a single item about presence of cognitive fluctuations (yes/no) and assigned a cognitive diagnosis (normal cognition, mild cognitive impairment [MCI] or dementia) at the baseline visit.^31^ Other variables were Montreal Cognitive Assessment (MoCA) score^32^ and a recently-developed and -utilized PPMI cognitive summary score (CSS) that is age- and sex-adjusted.^27,28^ For the scope of this study, MCI was defined as a MoCA score ≤ 25^33^ to be most consistent with rates of MCI reported in other iRBD cohorts, as a well-accepted cut-off for MCI,^33^ and as most likely to have clinical applicability (**Supplementary Table 1**).

#### Motor

Motor variables were MDS-UPDRS Part II total score and Part III total scores. In addition, subthreshold parkinsonism, the motor anchor for Stage 2 (i.e., subtle symptoms) in NSD-ISS, was defined as an MDS-UPDRS Part III score ≥ 5, excluding the postural and action tremor items.^30^

#### Neuropsychiatric

Neuropsychiatric variables were: (1) 15-item Geriatric Depression Scale (GDS-15)^34^; (2) State subscale of the State-Trait Anxiety Inventory (STAI)^35^; (3) MDS-UPDRS Part I Hallucination and Psychosis item score; and (4) MDS-UPDRS Part I Apathy item score. In addition, the use (yes/no) of three neuropsychiatric medication classes were recorded: benzodiazepine, antidepressant, and iRBD treatment (either clonazepam or melatonin).

#### Autonomic

Autonomic symptoms were presence of MDS-UPDRS Part I Urinary Problems score ≥ 1, Lightheadedness on Standing score ≥ 1, and Constipation Problems score ≥ 1.

#### Sensory

Sensory symptoms were presence of hyposmia (UPSIT score ≤ 15th percentile) and MDS-UPDRS Part I Pain and Other Sensations score ≥ 1.

#### Prodromal DLB criteria

As all individuals in the PSG-confirmed iRBD cohort had a confirmed proposed biomarker for prodromal DLB, only one additional core clinical feature beyond the essential criterion of MCI was needed.^7^ The core clinical features are fluctuating cognition, recurrent visual hallucinations, and parkinsonism; iRBD was not allowed to count as a core clinical feature as it was already an inclusion criterion for these analyses. MCI was diagnosed by MoCA score ≤ 25, presence of fluctuating cognition was a “yes” response by the LSI to the “cognitive fluctuations” question, recurrent visual hallucinations was MDS-UPDRS Part I Hallucination and Psychosis item score ≥ 1, and parkinsonism was at least “subthreshold” based on an MDS-UPDRS Part III Motor Examination score ≥ 5 (excluding postural or action tremor).

#### Neurobiological

Neurobiological variables were: (1) Amprion CSF neuronal α-synuclein SAA test results for determining presence of NSD^21^; (2) DaTscan results, reported as the mean striatal SBR and as dopaminergic dysfunction based on lowest putamen value < 75^th^ percentile SBR applying age-and sex-adjusted norms^26^; and (3) University of Pennsylvania Smell Identification Test (UPSIT) to assess olfaction.^36^ Hyposmia was defined as ≤ 15^th^ percentile applying age- and sex-adjusted norms for the revised UPSIT.^37,38^

### Statistics

Descriptive statistics at baseline were presented for iRBD participants and the robust HC group, including frequencies (percentages) for categorical measures, and means and standard deviations (SDs) or medians and interquartile ranges (IQRs) for continuous measures. Age- and sex-adjusted statistical comparisons of characteristics between the two groups were performed using logistic regression models for categorical measures, and general linear regression models for continuous measures. The Benjamini & Hochberg method was used to control the false discovery rate at 5%.

Descriptive statistics were also presented separately for the DaTscan-normal iRBD participants, DaTscan-abnormal iRBD participants, and robust HCs. Pairwise comparisons were performed between DAT-abnormal and DAT-normal iRBD participants using Wilcoxon rank sum tests for continuous measures, and Chi-square or Fisher’s exact test as appropriate for categorical measures. For comparisons between robust HCs, DAT-abnormal iRBD, and DAT-normal iRBD participants respectively, logistic regression models were used for categorical measures, and general linear regression models were used for continuous measures. These were adjusted for age and sex. Again, the Benjamini & Hochberg method was used to control the false discovery rate at 5%.

Criteria for probable prodromal DLB were evaluated for the overall iRBD cohort and were also reported by DaTscan status. The frequencies and percentages of participants who met each criterion were reported. MCI definitions were assessed by reporting their frequencies (percentages) across the iRBD NSD cohort.

The comorbid clinical symptoms of prodromal DLB among iRBD participants were assessed across five core clinical domains, and the frequency (percentage) of participants meeting the criteria for each was reported. An UpSet plot was used to display the percentage of participants with different combinations of comorbid clinical symptoms in the iRBD cohort. Prodromal LBD categories were reported based on the number of domains for which they met the criteria. The percentage of each clinical feature within each category was plotted on a cluster bar graph for participants with iRBD and one to four additional core clinical feature domains. The above analysis was repeated by applying more stringent criteria for defining the core clinical feature domains (i.e., for MDS-UPDRS Part I items having a score ≥ 2 for presence of the symptom). Odds ratios and 95% confidence intervals were reported for comparisons of the LBD symptoms between the iRBD group and the robust HC group.

Statistical analyses were performed using SAS v9.4 (SAS Institute Inc., Cary, NC; sas.com; RRID:SCR_008567). UpSet plots and bar charts were created using the “ComplexUpset” package in R Statistical Software (v 4.4.1; R Core Team 2025).

## RESULTS

### Characteristics of cohorts

#### Demographics

197 iRBD and 136 robust HC participants were included. iRBD participants were more likely to be male, white, Hispanic, and older, with a similar education level **(Table 1)**. There were 4 iRBD and 5 robust HC participants who had the non-Gaucher causing GBA1 genetic variants.

**Table 1.**
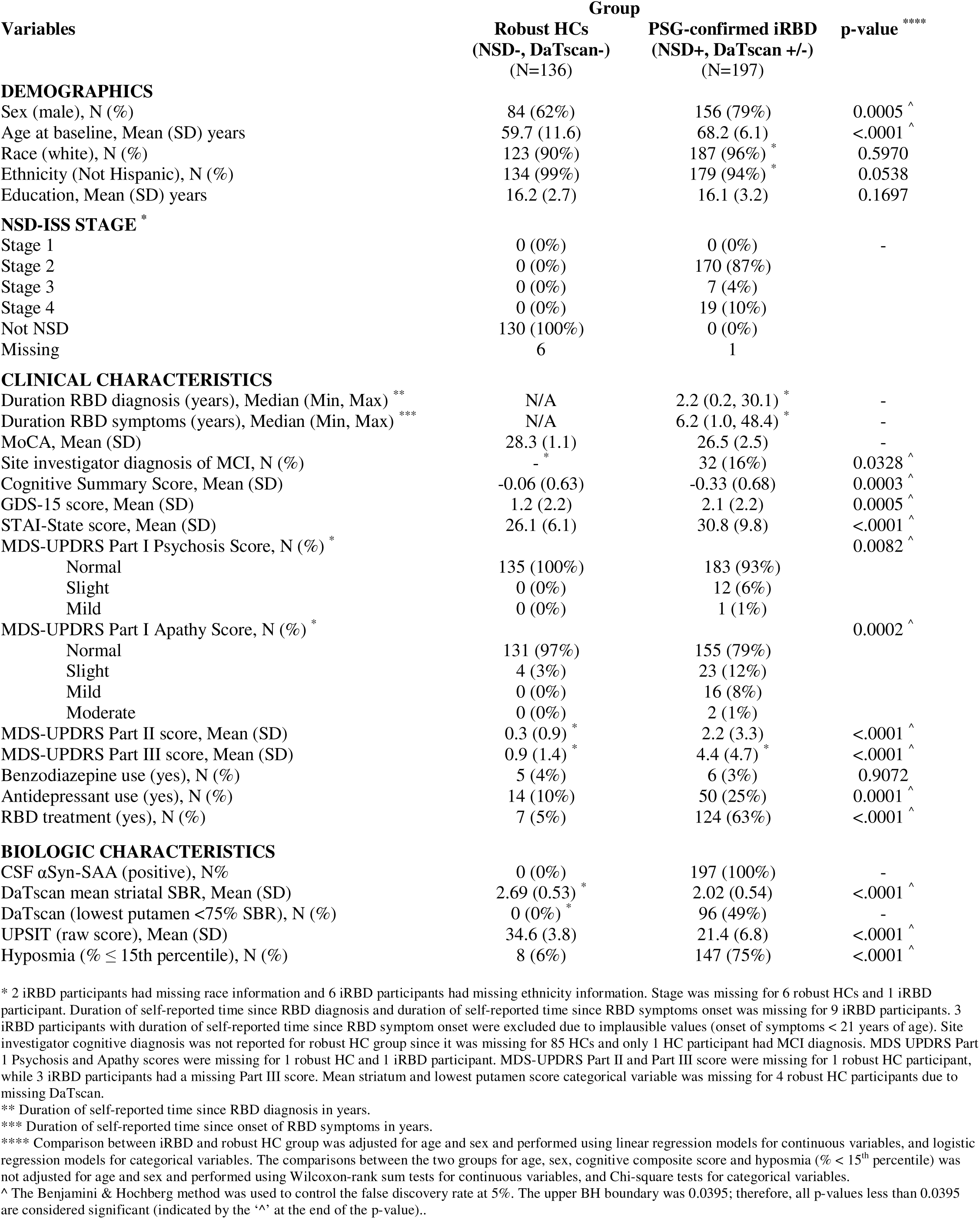
Characteristics of all NSD positive iRBD and robust HC participants.

| Variables | Group |  | p-value **** |
| --- | --- | --- | --- |
|  | Robust HCs<br>(NSD-, DaTscan-)<br>(N=136) | PSG-confirmed iRBD<br>(NSD+, DaTscan +/-)<br>(N=197) |  |
| DEMOGRAPHICS |  |  |  |
| Sex (male), N (%) | 84 (62%) | 156 (79%) | 0.0005 ^ |
| Age at baseline, Mean (SD) years | 59.7 (11.6) | 68.2 (6.1) | <.0001 ^ |
| Race (white), N (%) | 123 (90%) | 187 (96%) * | 0.5970 |
| Ethnicity (Not Hispanic), N (%) | 134 (99%) | 179 (94%) * | 0.0538 |
| Education, Mean (SD) years | 16.2 (2.7) | 16.1 (3.2) | 0.1697 |
| NSD-ISS STAGE * |  |  |  |
| Stage 1 | 0 (0%) | 0 (0%) | - |
| Stage 2 | 0 (0%) | 170 (87%) |  |
| Stage 3 | 0 (0%) | 7 (4%) |  |
| Stage 4 | 0 (0%) | 19 (10%) |  |
| Not NSD | 130 (100%) | 0 (0%) |  |
| Missing | 6 | 1 |  |
| CLINICAL CHARACTERISTICS |  |  |  |
| Duration RBD diagnosis (years), Median (Min, Max) ** | N/A | 2.2 (0.2, 30.1) * | - |
| Duration RBD symptoms (years), Median (Min, Max) *** | N/A | 6.2 (1.0, 48.4) * | - |
| MoCA, Mean (SD) | 28.3 (1.1) | 26.5 (2.5) | - |
| Site investigator diagnosis of MCI, N (%) | - * | 32 (16%) | 0.0328 ^ |
| Cognitive Summary Score, Mean (SD) | -0.06 (0.63) | -0.33 (0.68) | 0.0003 ^ |
| GDS-15 score, Mean (SD) | 1.2 (2.2) | 2.1 (2.2) | 0.0005 ^ |
| STAI-State score, Mean (SD) | 26.1 (6.1) | 30.8 (9.8) | <.0001 ^ |
| MDS-UPDRS Part I Psychosis Score, N (%) * |  |  | 0.0082 ^ |
| Normal | 135 (100%) | 183 (93%) |  |
| Slight | 0 (0%) | 12 (6%) |  |
| Mild | 0 (0%) | 1 (1%) |  |
| MDS-UPDRS Part I Apathy Score, N (%) * |  |  | 0.0002 ^ |
| Normal | 131 (97%) | 155 (79%) |  |
| Slight | 4 (3%) | 23 (12%) |  |
| Mild | 0 (0%) | 16 (8%) |  |
| Moderate | 0 (0%) | 2 (1%) |  |
| MDS-UPDRS Part II score, Mean (SD) | 0.3 (0.9) * | 2.2 (3.3) | <.0001 ^ |
| MDS-UPDRS Part III score, Mean (SD) | 0.9 (1.4) * | 4.4 (4.7) * | <.0001 ^ |
| Benzodiazepine use (yes), N (%) | 5 (4%) | 6 (3%) | 0.9072 |
| Antidepressant use (yes), N (%) | 14 (10%) | 50 (25%) | 0.0001 ^ |
| RBD treatment (yes), N (%) | 7 (5%) | 124 (63%) | <.0001 ^ |
| BIOLOGIC CHARACTERISTICS |  |  |  |
| CSF αSyn-SAA (positive), N% | 0 (0%) | 197 (100%) | - |
| DaTscan mean striatal SBR, Mean (SD) | 2.69 (0.53) * | 2.02 (0.54) | <.0001 ^ |
| DaTscan (lowest putamen <75% SBR), N (%) | 0 (0%) * | 96 (49%) | - |
| UPSIT (raw score), Mean (SD) | 34.6 (3.8) | 21.4 (6.8) | <.0001 ^ |
| Hyposmia (% ≤ 15th percentile), N (%) | 8 (6%) | 147 (75%) | <.0001 ^ |

\* 2 iRBD participants had missing race information and 6 iRBD participants had missing ethnicity information. Stage was missing for 6 robust HCs and 1 iRBD participant. Duration of self-reported time since RBD diagnosis and duration of self-reported time since RBD symptoms onset was missing for 9 iRBD participants. 3 iRBD participants with duration of self-reported time since RBD symptom onset were excluded due to implausible values (onset of symptoms < 21 years of age). Site investigator cognitive diagnosis was not reported for robust HC group since it was missing for 85 HCs and only 1 HC participant had MCI diagnosis. MDS UPDRS Part I Psychosis and Apathy scores were missing for 1 robust HC and 1 iRBD participant. MDS-UPDRS Part II and Part III score were missing for 1 robust HC participant, while 3 iRBD participants had a missing Part III score. Mean striatum and lowest putamen score categorical variable was missing for 4 robust HC participants due to missing DaTscan.
\*\* Duration of self-reported time since RBD diagnosis in years.
\*\*\* Duration of self-reported time since onset of RBD symptoms in years.
\*\*\*\* Comparison between iRBD and robust HC group was adjusted for age and sex and performed using linear regression models for continuous variables, and logistic regression models for categorical variables. The comparisons between the two groups for age, sex, cognitive composite score and hyposmia (% < 15<sup>th</sup> percentile) was not adjusted for age and sex and performed using Wilcoxon-rank sum tests for continuous variables, and Chi-square tests for categorical variables.
^ The Benjamini & Hochberg method was used to control the false discovery rate at 5%. The upper BH boundary was 0.0395; therefore, all p-values less than 0.0395 are considered significant (indicated by the '^' at the end of the p-value)..

#### NSD-ISS staging of iRBD participants

Approximately half (49%) were DaTscan abnormal (**Table 2**). Most (87%) iRBD participants were NSD-ISS Stage 2 at study enrollment; approximately 15% of participants were either Stage 3 (4%) or Stage 4 (10%). Of the 26 participants that were Stages 3 or 4, 16 met non-motor criteria, 10 cognitive criteria, and 4 motor criteria (4 participants met multiple criteria) (**Supplementary Table 2**).

**Table 2.**
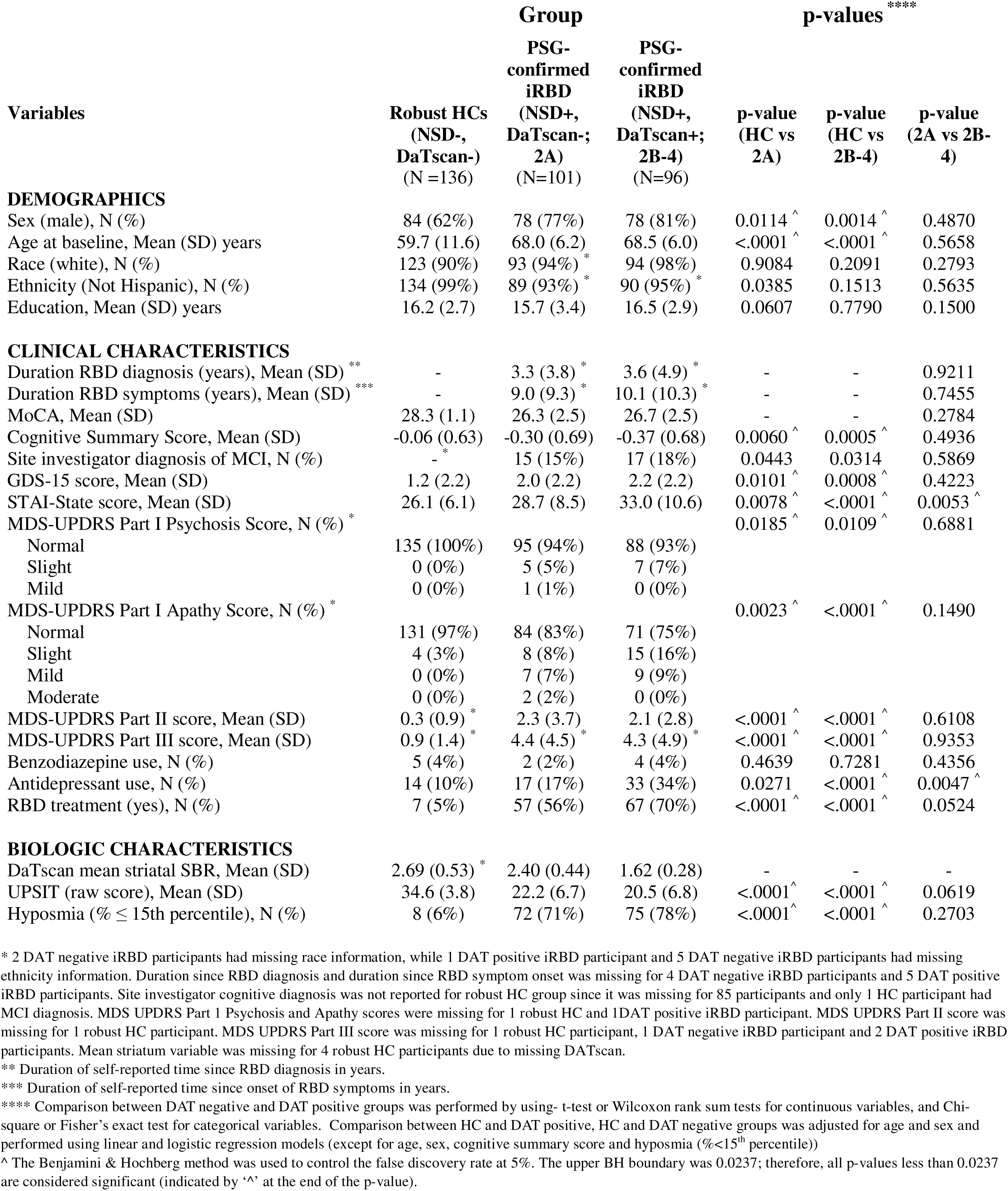
Characteristics of DAT positive iRBD, DAT negative iRBD and robust HC participants.

#### Clinical characteristics

The median duration of iRBD diagnosis was approximately two years, with median duration of symptoms being approximately six years (**Table 1**).

MoCA scores were approximately two points lower in the iRBD group. Nevertheless, their mean score remained within the unimpaired range, and only 16% of participants were diagnosed with MCI by the site investigator. The CSS was significantly lower in the iRBD group (*z*-score difference = 0.27, *p*-value = 0.0003, effect size = 0.41).

Motor scores (MDS-UPDRS Parts II and III) were higher in iRBD than in HCs but were low overall.

The iRBD group had higher depression and anxiety scores on disorder-specific rating scales than did the HC group, and using the cut-off scores on the MDS-UPDRS Part I had higher rates of psychosis and apathy. Applying cut-off scores to all neuropsychiatric symptoms within the iRBD group, the rates of depression, anxiety, and apathy were all higher than psychosis.

There was a strong association between iRBD status and olfaction, with 75% of iRBD participants having hyposmia versus 6% of HCs.

Approximately 2/3 of iRBD participants were taking a medication for their sleep disorder, and 1/4 were on an antidepressant. We found no association between sleep medication use and either the MoCA (*p*-value = 0.6946) or the CSS (*p*-value = 0.2411).

#### Impact of dopamine system dysfunction in iRBD participants

Demographic characteristics and iRBD duration were similar for Stages 2A and 2B participants (**Table 2**). There was no impact of an abnormal DaTscan on cognitive performance as assessed with the MoCA, CSS, or LSI cognitive diagnosis. Likewise, there were no between-group differences in motor scores. For neuropsychiatric symptoms, anxiety scores were higher in those iRBD participants with an abnormal DaTscan. Antidepressant use was more prevalent in Stage 2B.

#### Criteria for probable prodromal Lewy body dementia

After applying the prodromal DLB criteria,^9^ only 10% of iRBD participants met the criteria (**Supplementary Table 3**). Even though approximately one-third (32%) of iRBD participants met the criterion for MCI, only 27% of the entire cohort met the criterion for subthreshold parkinsonism, 7% for psychosis, and 2% for fluctuating cognition. The results were similar for those in Stage 2A versus 2B.

#### Prevalence of symptoms across five clinical domains in iRBD

By moving beyond existing diagnostic criteria and evaluating symptoms across a broader range of clinical domains known to be affected in prodromal Lewy body disease, we found that individuals with iRBD exhibit a diverse spectrum of impairments.

Applying less stringent cut-offs for MDS-UPDRS items (i.e., score ≥ 1 for presence of symptom), rates of any sensory symptom, any autonomic symptom, and any neuropsychiatric symptom all exceeded 50%, higher than rates for MCI or subthreshold parkinsonism (**Table 3**, **Figure 1**).

**Figure 1.**
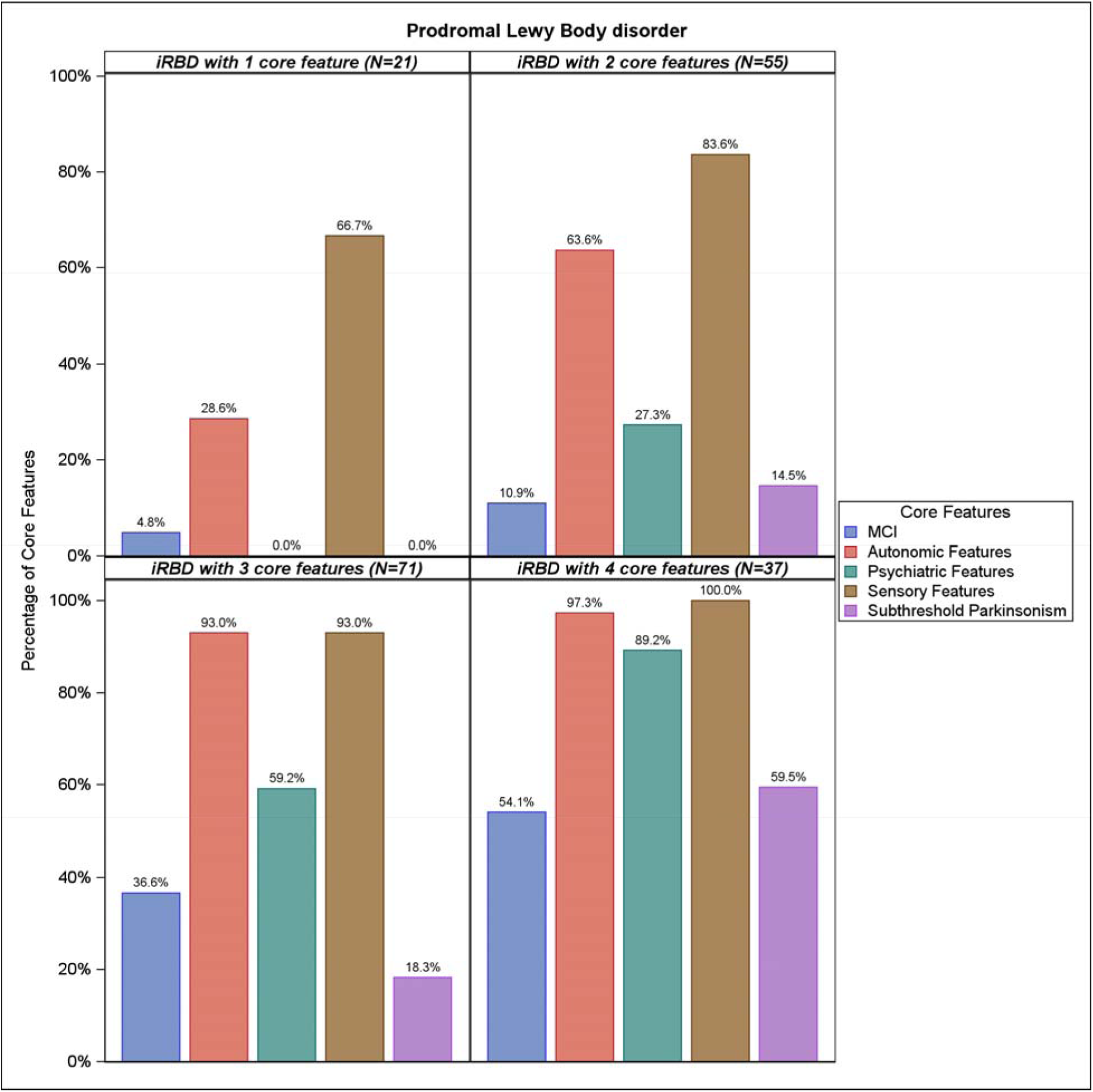
Bar graphs of frequency of comorbid clinical domains in iRBD using original criteria.

**Table 3.**
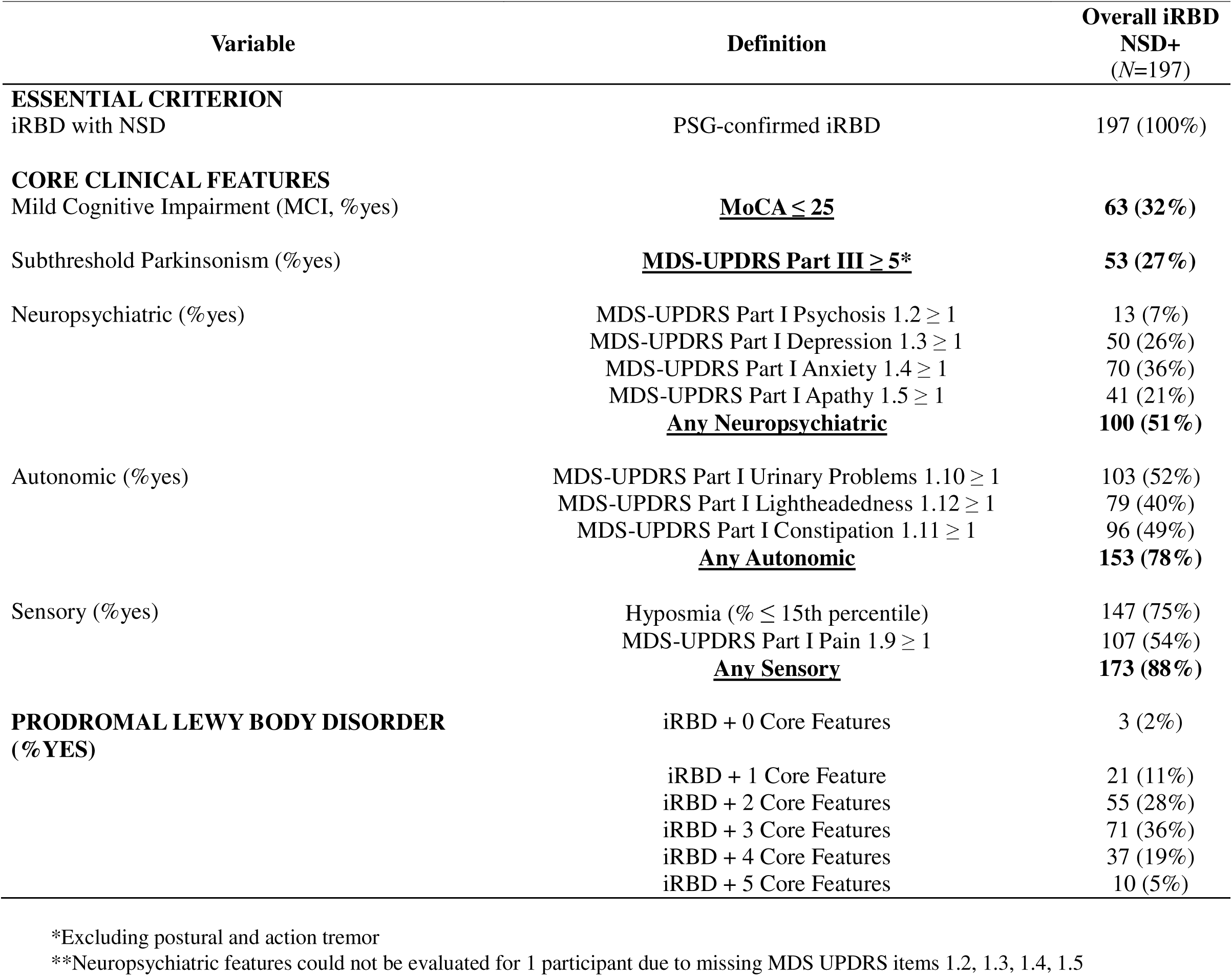
Comorbid clinical symptoms of prodromal Lewy body disorder in iRBD with synucleinopathy.

Applying more stringent cut-offs for MDS-UPDRS items, rates of sensory impairment remained very high (due to the criterion for hyposmia being unchanged), and autonomic and neuropsychiatric symptoms were of comparable prevalence to MCI and parkinsonism (**Supplementary Table 4**, **Supplementary Figure 1**).

**Figure 2** is an UpSet plot based on less stringent MDS-UPDRS cut-offs, and there were nine additional symptoms or combination of symptoms with ≥ 5% prevalence, together representing 79% of the entire cohort. **Supplementary Figure 2** is an UpSet plot based on more stringent MDS-UPDRS cut-offs.

**Figure 2.**
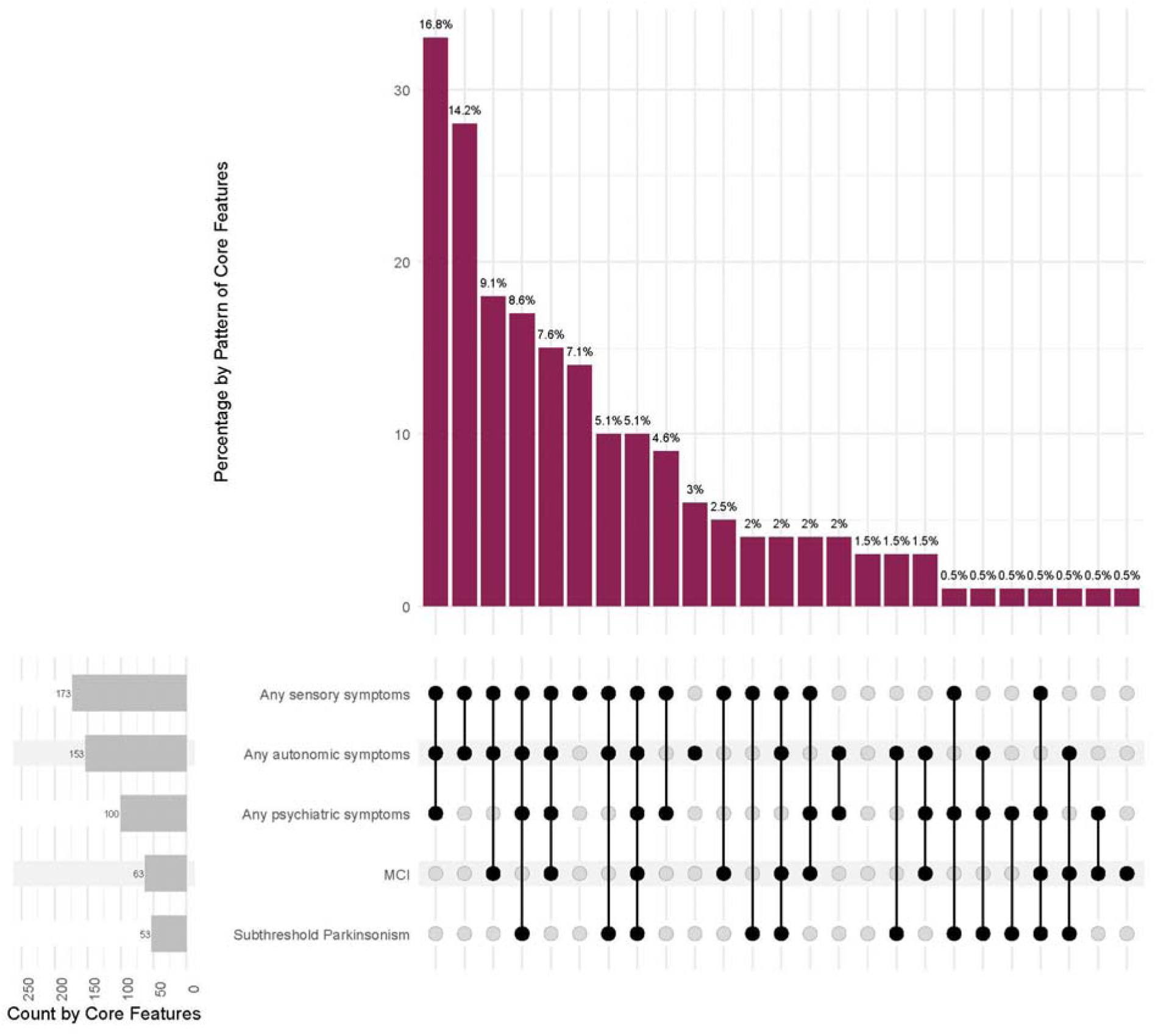
UpSet plot of overlap of comorbid clinical features.

#### Odds of domain impairment in iRBD versus healthy controls

Symptoms in all four non-cognitive domains were significantly more common in the iRBD cohort compared with HCs, regardless of less or more stringent MDS-UPDRS cut-offs being applied (**Supplementary Table 5**). Although the prevalence of sensory, autonomic, and neuropsychiatric symptoms decreased among iRBD participants when more stringent cut-offs were applied, the odds ratios for the prevalence of these symptoms in iRBD compared with HCs increased. The highest odds ratios were for parkinsonism and sensory symptoms. We reran the odds ratios for iRBD versus HCs adjusting for age and sex, and the results were similar (**Supplementary Table 6**).

## DISCUSSION

In a multi-center, international cohort, participants with iRBD and *in vivo* biomarker-defined synucleinopathy, but without a clinically-diagnosed neurodegenerative disease, demonstrated poorer performance than a robust healthy control group across multiple clinical domains, including cognitive, motor, neuropsychiatric, autonomic, and sensory. Typically, multiple-domain impairment combinations were observed. Clinical associations of dopaminergic deficiency as an additional biological feature were observed, notably greater neuropsychiatric comorbidity, but not parkinsonism or cognition. The most common individual symptoms or features in the iRBD group were hyposmia, pain and other sensations, urinary dysfunction, constipation, lightheadedness on standing, and anxiety, whereas the core clinical features of LBDs, parkinsonism, and cognitive impairment were less common. Importantly for clinical research, we also found that existing research diagnostic criteria for prodromal LBDs^9^ may be too restrictive or fail to capture certain symptoms that are commonly observed in the early stage of synuclein disease.

Our results indicate that cognitive deficits are present in iRBD regardless of dopaminergic deficit. This suggests that the neurobiological basis for early cognitive decline in this population is in part non-dopaminergic and non-striatal, consistent with cholinergic^39^ and noradrenergic^40,41^ deficits present in iRBD, as well as Lewy body pathology observed postmortem in extra-striatal regions (e.g., brainstem, limbic system), even in individuals without a neurodegenerative disease diagnosis.^42^ We found that having a positive DaTscan was associated with higher anxiety scores and antidepressant use, which is consistent with previous research showing a role for the dopamine system in affective symptoms in PD,^43^ or perhaps reflecting the involvement of other brainstem nuclei (e.g., raphe) associated with affective symptoms.^44,45^

One objective of our research was to determine how many iRBD participants met the research diagnostic criteria for prodromal DLB.^9^ Regarding the essential clinical feature of MCI, we found that approximately 1/3 of iRBD participants met the criterion based on the MoCA cut-off score and that their impairment on the CSS was of a medium effect size, which is consistent with previous research findings of high rates of cognitive deficits in iRBD.^10,11^ However, the other core clinical features were uncommon. Thus, formal prodromal DLB criteria was met in only 10% of iRBD participants, indicating that this prodromal syndrome, as currently defined, is uncommon in recently-diagnosed iRBD.

Moving beyond existing diagnostic criteria revealed a substantially more complex symptom profile. All five clinical domains examined were more impaired in iRBD than in the robust HC group, with the highest rate found for the sensory domain due to almost 75% of iRBD participants having hyposmia, which has been reported to be more common in iRBD than in healthy controls.^18^ In addition, persons with comorbid iRBD and hyposmia have been reported to have elevated rates of cognitive deficits, motor symptoms, depression, and apathy.^46^ In addition, elevated rates of neuropsychiatric^47^ and autonomic^48^ symptoms have also been demonstrated in iRBD.

Following hyposmia, the most frequently-reported symptoms included another sensory symptom (pain), autonomic symptoms such as urinary dysfunction, constipation, and lightheadedness, as well as neuropsychiatric symptoms, primarily anxiety. In contrast, the diagnostic signs and symptoms of PD (motor) and DLB (dementia) were less prevalent, as were fluctuating cognition and psychosis. We also found that multi-clinical domain impairment was common, and in many combinations. This significant co-morbidity was despite the median duration of iRBD being only two years, which is notably shorter at study entry than previously-reported cohorts.^49–51^

Limitations of the research worth noting are having only single items for certain symptoms (e.g., cognitive fluctuations, apathy and psychosis), defining MCI on the basis of a MoCA cut-off score, and the presence of demographic differences (i.e., age, sex, race and ethnicity) between the iRBD and HC groups. Regarding the latter point, the odds ratios for frequency of clinical domains affected in the iRBD versus the HC groups were similar when adjusted for age and sex, and the cognitive summary score utilized is age- and sex-corrected. Also, we chose to focus only on probable DLB criteria, not possible DLB. In addition, we chose to focus on iRBD participants who had biomarker-defined synucleinopathy, so we did not compare those iRBD participants who are α-synuclein SAA+ versus those who are SAA-, which can be done in future studies.

Although PD and DLB are diagnosed by the presence of parkinsonism and dementia, our results show that in iRBD with biologically-defined synucleinopathy, clinical features in other domains (i.e., neuropsychiatric, autonomic, and sensory) are equally or more prevalent, suggesting that widespread synuclein pathology is present even in recently-diagnosed iRBD. Moreover, symptoms across all clinical domains occur far more frequently in iRBD than in healthy controls and co-occur in diverse combinations. These findings highlight the importance of assessing multiple clinical domains in early or prodromal synuclein disease rather than anchoring diagnostic criteria or assessment instruments solely to motor symptoms or cognitive impairment.

## ACKNOWLEDGEMENTS

PPMI – a public-private partnership – is funded by the Michael J. Fox Foundation for Parkinson’s Research and funding partners, including 4D Pharma, Abbvie, AcureX, Allergan, Amathus Therapeutics, Aligning Science Across Parkinson’s, AskBio, Avid Radiopharmaceuticals, BIAL, BioArctic, Biogen, Biohaven, BioLegend, BlueRock Therapeutics, Bristol-Myers Squibb, Calico Labs, Capsida Biotherapeutics, Celgene, Cerevel Therapeutics, Coave Therapeutics, DaCapo Brainscience, Denali, Edmond J. Safra Foundation, Eli Lilly, Gain Therapeutics, GE HealthCare, Genentech, GSK, Golub Capital, Handl Therapeutics, Insitro, Jazz Pharmaceuticals, Johnson & Johnson Innovative Medicine, Lundbeck, Merck, Meso Scale Discovery, Mission Therapeutics, Neurocrine Biosciences, Neuron23, Neuropore, Pfizer, Piramal, Prevail Therapeutics, Roche, Sanofi, Servier, Sun Pharma Advanced Research Company, Takeda, Teva, UCB, Vanqua Bio, Verily, Voyager Therapeutics, the Weston Family Foundation and Yumanity Therapeutics.

## AUTHOR CONTRIBUTIONS

Conception and design of the study: (author initials)

Acquisition and analysis of data:

Drafting of the manuscript and figures:

Contributions from the PPMI study group can be found in **Supplementary File 1.**

## DATA AVAILABILITY

Data used in the preparation of this article were obtained on Jan 21, 2025, from the Parkinson’s Progression Markers Initiative (PPMI) database (www.ppmi-info.org/access-data-specimens/download-data), RRID:SCR_ 006431. For up-to-date information on the study, visit www.ppmi-info.org.

This analysis was conducted by the PPMI Statistics Core and used actual dates of activity for participants, a restricted data element not available to public users of PPMI data. This analysis used DAT-SPECT and aSynSAA results for participants of the Prodromal Cohort, obtained from PPMI upon request after approval by the PPMI Data Access Committee.

Protocol information for The Parkinson’s Progression Markers Initiative (PPMI) Clinical - Establishing a Deeply Phenotyped PD Cohort AM 3.2. can be found on protocols.io or by following this link: https://dx.doi.org/10.17504/protocols.io.n92ldmw6ol5b/v2.

Statistical analysis codes used to perform the analyses in this article are shared on Zenodo [10.5281/zenodo.17611026].

